# Quality assessment of endotoxin contamination in consumables used for assisted reproductive technology

**DOI:** 10.64898/2026.01.06.26343566

**Authors:** Hiroyuki Tomari, Emi Sugizaki, Shaimaa Ibrahim, Yuki Hashiguchi, Genki Koyama, Yasuo Nakamura, Mayu Nagata, Yumi Nagata, Yuji Haishima

**Author notes:** Corresponding author: (YH). These authors contributed equally to this paper (Hiroyuki Tomari, Emi Sugizaki).

## Abstract

Commercially available disposable products such as cell culture utensils and catheters do not necessarily possess sufficient quality for in vitro fertilization (IVF), from the perspective of pyrogen contamination. We aimed to comprehensively analyze the pyrogen contamination status, including bacterial endotoxins, of the products used for IVF in assisted reproductive technology (ART) and improve their cleanliness using a new sterilization technology. Pyrogen contamination levels were evaluated using a direct human cell-based pyrogen test that is not affected by the recovery ratio, unlike bacterial endotoxin tests. Pyrogen inactivation tests were performed using low-temperature ozone/hydrogen peroxide gas treatment. The residual hydrogen peroxide was colorimetrically quantified, and the effectiveness of its removal by drying treatment was evaluated using germ cell viability as an indicator. Significant amounts of pyrogen, from 0.014 to 1.110 EU/product, were detected in seven of the twenty products. Pyrogen contamination levels were reduced below the detection limit by ozone/hydrogen peroxide gas sterilization. Hydrogen peroxide remained on the surface of the GPS dish but was reduced to a level that did not affect human sperm viability and embryo development after drying at 80°C for 24 h following sterilization. These products may carry a potential risk of reducing ART success rates, and pyrogen contamination levels may exceed the previously reported allowable level of 0.01–0.02 EU/mL in human IVF during actual use. Our study suggests that the manufacturing of products free from pyrogens and without adverse effects on germ cells is possible using ozone/hydrogen peroxide gas sterilization and subsequent drying technologies.

## Introduction

Assisted reproductive technology (ART) is any treatment or method that involves the handling of human oocytes, sperm, or embryos in order to achieve pregnancy. In general, it is a generic term for infertility treatment methods such as in vitro fertilization (IVF) and embryo transfer, intracytoplasmic sperm injection and embryo transfer, and frozen-thawed embryo transfer [1].

The physical and chemical effects, such as the quality of the culture medium and utensils, the culture conditions (temperature, humidity, and vapor phase), and the presence or absence of infection, are important determinants of the safety and success rate of ART [2,3]. It has been proposed that the most important of these effects may be the quality of the culture medium, in particular the level of endotoxin contamination, which is the most potent pyrogen currently known and exhibits a broad spectrum of biological activities in vitro and in vivo, even at low concentrations [4,5]. Snyman and Van der Merwe demonstrated that medium contaminated with endotoxin levels greater than 1 ng/mL reduced pregnancy rates in human IVF programs and reported that endotoxins cause embryo fragmentation [6]. Nagata et al. reported that the standard for the endotoxin levels in culture medium should be less than 1 pg/mL to obtain the best outcome, and the acceptable level should be set to 2 pg/mL in human IVF [2,3]. In an experiment using an in vitro rat embryo test system with water of different qualities, Han Q et al. showed that the group using super-purified water without endotoxin and nucleic acid had the best results in terms of embryo formation rate, total cell number, and inner cell number [7].

Studies on the effects of endotoxin on sperm have not yielded uniform results. Dumoulin JC et al. reported that human sperm viability and the IVF of mouse oocytes, followed by subsequent zygote culture, were unaffected by relatively high endotoxin concentrations [8]. However, subsequent studies have reported that sperm expressing Toll-like receptors [9] on their membranes can recognize endotoxins, resulting in significant impairment of fertilization potential due to reduced motility and increased apoptosis [10,11]. In addition, sperm may be highly sensitive to endotoxin because the epithelium of the human epididymis expresses the gene encoding lipopolysaccharide-binding protein (LBP), which plays an important role in mediating the biological activity of endotoxin [12]. LBP was also detected in cells and attached to the heads and tails of spermatozoa [13]. A potential relationship between endotoxin levels in amniotic fluid and the onset of preterm labor has also been reported [14]. Viana GA reported a high incidence of endometritis in infertile couples, with bacterial endotoxin levels in menstrual samples higher in patients with suspected endometritis [15].

Thus, endotoxin significantly influences the outcome of a human IVF-embryo transfer program. With improvements in water quality and the widespread use of prepared products, problems related to endotoxin contamination in culture medium are being resolved. On the other hand, few studies have evaluated the quality of commonly used cell culture utensils distributed as pyrogen-free products. We recently found that commercially available cell culture utensils were not necessarily pyrogen-free, and some products were contaminated with pyrogen exceeding the acceptable level [16], using a direct human cell-based pyrogen assay (HCPT) that is not affected by recovery ratio, unlike the endotoxin test [17,18]. This finding indicates that products contaminated with pyrogen may also exist among culture utensils used for ART. In this study, to confirm and address this issue, we conducted a comprehensive investigation of pyrogen contamination in currently available utensils commonly used in ART and verified the pyrogen inactivation effects of ozone/hydrogen peroxide gas sterilization, which is the first method to effectively sterilize and simultaneously decompose organic compounds, including endotoxin, at 50℃ [19,20]. In addition, we established a method to remove residual hydrogen peroxide on the surface after sterilization and evaluated its impact on human sperm viability and embryo development.

## Materials and methods

### Ethics statement

The protocols of laboratory experiments with human-derived samples used in this study were approved by the Ethical Committee of Miura Co., Ltd. and IVF Nagata Clinic (approval numbers 20240822-1 and 201130-2) and complied with the Helsinki Declaration of 1964 and its later amendments. Informed consent was obtained from all patients for being included in the study. Blood donors for HCPT and donors of human sperm and embryo were recruited after obtaining research ethics approval. Written informed consent was obtained from each donor after explaining the details of the research content with related documents. Animal care or treatments were in agreement with the Animal Care and Use Protocol of Astec Cell Science Institute, Fukuoka, Japan.

### Data availability

The data that support the findings of this study are fully available without restriction. Disclosure of name and Lot number of products applied to HCPT are also available from the corresponding author upon reasonable request.

### Test sample

Test samples used in this study are listed in Table 1. All samples were randomly extracted from materials routinely used for IVF at Nagata Clinic. These hospital-sourced samples were selected to reflect real-world clinical practice. The samples were purchased from the manufacturers of each product. Detailed information about specific products, including company names and catalog numbers, is available upon reasonable request.

**Table 1.** Samples and the conditions of direct HCPT used for surveying the contamination status of pyrogen.

| Sample no. | Product type | Amount of culture medium (mL) | Number of samples | Culture vessel | Quality labelling |
| --- | --- | --- | --- | --- | --- |
| 1 | Culture dish | 2 | 3 | Direct | Sterile, < 20 EU/device |
| 2 | Culture dish | 3 | 3 | Direct | Sterile, < 20 EU/device |
| 3 | Culture dish | 2 | 3 | Direct | Sterile |
| 4 | Culture dish | 5 | 3 | Direct | Sterile, < 0.03 EU/device |
| 5 | Dropper / Pasteur pipette | 150 | 5 | Cell culture flask<sup>2)</sup> | Sterile, non-pyrogenic |
| 6 | Dropper / Pasteur pipette | 150 | 5 | Cell culture flask | Sterile, < 0.25 EU/device |
| 7 | 15mL conical tube | 2 | 3 | Direct | Sterile, < 0.5 EU/device |
| 8 | 15mL conical tube | 1 | 3 | Direct | Sterile, < 0.03 EU/device |
| 9 | 15mL conical tube | 1 | 3 | Direct | Sterile, non-pyrogenic |
| 10 | 1mL microchip | 150 | 50 | Cell culture flask | Sterile, DNA & Nuclease free |
| 11 | Center well dish | 1 | 3 | Direct | Sterile, < 0.03 EU/device |
| 12 | Centrifuge tube | 150 | 5 | Cell culture flask | Sterile |
| 13 | 5mL round tube | 1 | 3 | Direct | Sterile, non-pyrogenic |
| 14 | Serum vial | 2 | 3 | Direct | Sterile |
| 15 | Intra-uterine insemination catheter | 150 | 3 | Cell culture flask | Medical device |
| 16 | Intra-uterine insemination catheter | 150 | 3 | Cell culture flask | Medical device |
| 17 | Embryo transfer catheter | 150 | 3 | Cell culture flask | Medical device |
| 18 | Embryo transfer catheter | 150 | 3 | Cell culture flask | Medical device |
| 19 | Embryo transfer catheter | 150 | 3 | Cell culture flask | Medical device |
| 20 | Embryo transfer catheter | 150 | 3 | Cell culture flask | Medical device |

Note: Direct: culture medium was directly added to each product. 2) Cell culture flask: all samples were immersed together in a single flask (base area 150 cm<sup>2</sup>).
EU, endotoxin unit; HCPT, human cell-based pyrogen test.

### Direct HCPT

Human blood was obtained from a volunteer at our laboratory and immediately mixed with heparin (10 units/mL). The procedure was performed in accordance with the ethical guidelines of MIURA CO., LTD. (approval numbers: 20231213-1 and 20240822-1). To prepare the culture medium, the heparinized blood was diluted tenfold with Roswell Park Memorial Institute Medium, formulation 1640, containing fetal bovine serum (10%), glutamine (2 mM), non-essential amino acids (0.1 mM), sodium pyruvate (1 mM), and bovine insulin (9 µg/mL).

HCPT was performed according to previously reported methods [16,19,20]. Briefly, an appropriate number of sample nos. 5, 6, 10, 12, and 15 to 19 were soaked in a cell culture flask (bottom area: 150 cm^2^) containing 150 mL of the culture medium (Table 1) and incubated for 17 h at 37°C under a 5% CO_2_ atmosphere. Suitable amounts of the medium were directly added to the other samples (Table 1) and cultured under the same conditions.

The culture supernatants of each sample were collected individually after centrifugation and stored at -30℃ until use. Interleukin-6 (IL-6) levels in the supernatants were measured using the Human IL-6 Quantikine ELISA kit purchased from R&D Systems (Minneapolis, MN, USA). The IL-6 concentrations (pg/mL) in each supernatant were converted to endotoxin units (EUs)/mL using a standard curve prepared with several concentrations of Japanese Pharmacopoeia Endotoxin Reference Standard, purchased from the Pharmaceutical and Medical Device Regulatory Science Society of Japan (Tokyo, Japan). Limit of detection (LOD) and limit of quantification (LOQ) were calculated by adding 3 or 10 times the standard deviation to the background mean. The mean values of the LOD and LOQ in all experiments were 0.0025 and 0.0048 EU/mL, respectively.

Total pyrogen amounts of samples were calculated by multiplying the volume of culture medium used in HCPT (Table 1) by the pyrogen concentrations (EU/mL) determined by ELISA. In Table 2, for sample nos. 5, 12, and 16 to 18, “Max” represents the worst-case scenario, where the total pyrogen is assumed to be present in only one of the samples used for HCPT. “Min” represents the pyrogen level, assuming uniform contamination across all samples used for HCPT. In cases of sample nos. 7 and 14, “Max” and “Min” correspond to the highest and lowest pyrogen contamination detected among the samples applied to HCPT, respectively.

**Table 2.**
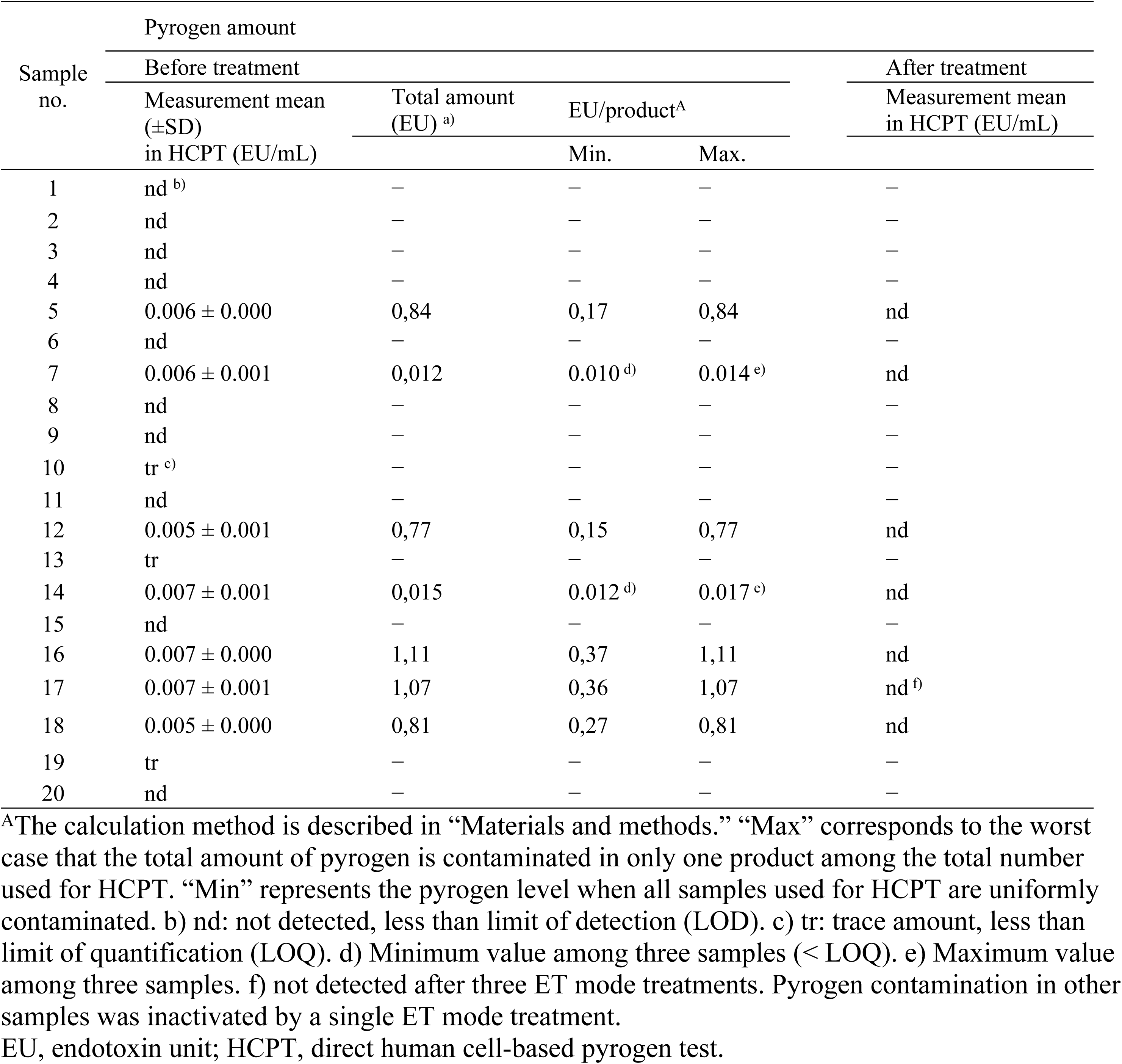
Pyrogen amount detected from samples before and after ozone/hydroxy peroxide gas treatment in elaborate treatment (ET) mode.

### Ozone/hydrogen peroxide gas sterilization and removal of residual hydrogen peroxide

Ozone/hydrogen peroxide gas sterilization of utensils sealed in sterile bags was performed with XZ-100 (MIURA CO., LTD., Ehime, Japan) in elaborate treatment (ET) mode for endotoxin inactivation (Ibrahim *et al*. 2025) at 50℃. µDrop GPS dish (Lifeglobal Group, LCC, CT, USA) used for the human sperm viability test and embryo assay described below was treated by different methods as shown in Table 3. For test groups A and B, the dishes were treated with the default ET mode setting (aeration: 5 cycles) or with a modified setting in which the number of aeration cycles was increased to 455 to enhance hydrogen peroxide removal. For test groups C to E, the dishes sterilized by XZ-100 in ET mode with the default setting were dried using an RL dryer (MIURA CO., LTD., Ehime, Japan) for 3, 5, and 24 h at 80℃ instead of aeration in XZ-100.

**Table 3.** Effect of aeration on removal of residual hydrogen peroxide.

| Group | Sterilization with XZ100 (mode / number of aeration cycles) | Aeration with RL dryer at $80^\circ\text{C}$ (hr) | Amount of residual hydrogen peroxide ($\mu\text{g/mL}$)<sup>A</sup> |
| --- | --- | --- | --- |
| Control | — | — | — |
| A | ET / 5 | — | $123.34 \pm 5.30$ |
| B | ET / 455 | — | $0.37 \pm 0.03$ |
| C | ET / 5 | 3 | $2.65 \pm 0.21$ |
| D | ET / 5 | 5 | $0.60 \pm 0.10$ |
| E | ET / 5 | 24 | $0.14 \pm 0.02$ |

Note: Default setting for the sterilization in elaborate treatment (ET) mode. Values are the mean ± standard deviation.
<sup>A</sup>The estimated concentration when human sperm was cultured in the center well of μDrop GPS dish with 2 mL of medium. The concentration during human embryo culture is 1.5 times lower based on the amount of medium used.

### Quantification of hydrogen peroxide

Two milliliters of purified water, equal to the volume of culture medium normally used for sperm culture, was dispensed into the center well of the µDrop GPS dish treated under different conditions described above, and hydrogen peroxide was extracted for 24 h at room temperature. The hydrogen peroxide concentration of each extract was measured using a SpectroQuant Hydrogen Peroxide Test Kit (Merck Millipore, Billerica, MA, USA) and a microplate reader SH-9000Lab (Hitachi High-Tech Science Corporation, Tokyo, Japan) at a wavelength of 445 nm (Ibrahim *et al*. 2025). The LOD was 0.015 µg/mL, as specified by the catalog.

### Embryo assay

The mouse embryo assay was performed at an external laboratory (Astec Cell Science Institute, Fukuoka, Japan) using frozen pronuclear embryos derived from BDF1 mice. Culture dishes were prepared by adding 20 μL of Potassium Simplex Optimized Medium (ARK Resource Co., Ltd., Kumamoto, Japan) to each well of a µDrop GPS dish and overlaying 2.5 mL of mineral oil (M5310, Merck, Tokyo, Japan). Twenty-one frozen pronuclear embryos were seeded nearly evenly into five wells per dish and cultured for 96 h at 37°C in a 5% CO_2_ atmosphere. The blastocyst formation rate was evaluated after 96 h of culture. The control group, which included the commercially available product without modification, was compared with the test groups B, E, and F, as presented in Table 3.

For human embryo assays (HEAs), 139 frozen human 4-cell stage (day 2) embryos from patients who consented to research use and disposal were used. The procedure was performed in accordance with the ethical guidelines of IVF Nagata Clinic (approval number: 201130-2, including the sperm survival test described below). Culture dishes were prepared by adding 30 μL of GLOBAL medium (Lifeglobal Group, CT, USA) to each well of a µDrop GPS dish and overlaying 3.0 mL of mineral oil (Fuso Pharmaceutical Industries, Ltd., Osaka, Japan). One 4-cell embryo was seeded in each well and cultured at 37℃ in 5% CO_2_ and 5% O_2_ until day five (3 days). The blastocyst rate was evaluated by observing blastocoel formation using a time-lapse incubator (CCM-iBIS, Astec Co., Ltd., Fukuoka, Japan). In the HEA, full blastocysts and good blastocysts (3BB or higher) were also evaluated using the Gardner classification [21]. The control group, which included the commercially available product without modification, was compared with the test groups B and F, as presented in Table 3.

All experiments were performed in duplicate for each treatment group, and the mean values were used for subsequent analyses.

### Sperm survival test

A sperm survival test was conducted using discarded semen from nine consenting patients. Well-motile human sperm were separated using a density gradient solution (ORIGIO® Gradient™, CooperSurgical Inc., CT, USA), and sperm motility was adjusted to over 90% using the swim-up method. A total of 100 μL of the adjusted sample was seeded into a µDrop GPS dish and cultured with 2.0 mL of human tubal fluid medium (Lifeglobal Group, CT, USA) for 48 h at 37°C under 5% CO_2_ and 5% O_2_. Sperm motility was assessed immediately after preparation and after 24 and 48 h of incubation at 37°C. The percentage of motile sperm was evaluated by counting moving and non-moving spermatozoa. Sperm motility was observed using an inverted microscope (IX71, Olympus, Tokyo, Japan) at 100x magnification, and at least 200 sperm were counted in three fields of view. The control group, which included the commercially available product without modification, was compared with the test groups B and F, as presented in Table 3.

### Statistical analysis

Differences between groups in the proportion of results were evaluated using the Student’s t-test. Statistical analyses were performed using EZR (Saitama Medical Center, Jichi Medical University, Saitama, Japan). A p-value <0.05 was considered statistically significant.

## Results

### Pyrogen contamination survey of utensils used for human IVF

The pyrogen contamination level of the currently available commercial utensils for human IVF was evaluated by direct HCPT. As shown in Table 2, sample nos. 1 to 4, 6, 8, 9, 11, 15, and 20 were not contaminated with pyrogen, with measurement values below the LOD. However, pyrogen of 0.005 to 0.007 EU/mL exceeding LOQ was detected from nos. 5, 7, 12, 14, and 16 to 18. The total amounts of pyrogen corresponded to 0.84, 0.012, 0.77, 0.015, 1.11, 1.07, and 0.81 EU, respectively, based on the volume of culture medium used (Table 1). The amounts of pyrogen, when all tested pieces of nos. 5, 12, and 16 to 18 (Table 1) were uniformly contaminated, were calculated as 0.17, 0.15, 0.37, 0.36, and 0.27 EU/product, while the worst cases, assuming the detected pyrogens were contaminated in one product alone, were 0.84, 0.77, 1.11, 1.07, and 0.81 EU/product, respectively. Minimum and maximum amounts of pyrogen detected from three pieces of no. 7 were 0.010 and 0.014, and those detected from no. 14 were 0.012 and 0.017 EU/product, respectively. In addition to these seven samples, trace amounts of pyrogen, less than LOQ and more than LOD, were detected from nos. 10, 13, and 19.

### Effect of ozone/hydrogen peroxide gas treatment for depyrogenation

Ozone/hydrogen peroxide gas treatment with ET mode was applied to clearly pyrogen-positive samples (nos. 5, 7, 12, 14, 16, 17, and 18) for evaluating the depyrogenation ability. As shown in Table 2, no pyrogen above LOD was detected from a product after the treatment.

### Removal performance of residual hydrogen peroxide by drying treatment

The µDrop GPS dish was selected as a representative utensil for human sperm and embryo culture. As shown in Table 3, 123.34±5.30 µg/mL of hydrogen peroxide remained on the surface of the dish after ozone/hydrogen peroxide gas treatment followed by five aeration cycles in the same sterilizer (test group A). The amount was reduced to 0.37±0.03 µg/mL by increasing the number of aeration cycles to 455 (test group B). Instead of aeration in the sterilizer, drying treatment at 80℃ was also effective in reducing residual hydrogen peroxide to 2.65±0.21, 0.60±0.10, and 0.14±0.02 µg/mL after 3, 5, and 24 h of treatment (test groups C to E).

### Effect on mice and human embryo development

To minimize the sample number from the perspective of animal welfare and effective utilization of human-derived materials, the evaluation of residual hydrogen peroxide effects on germ cells was confined to the µDrop GPS dishes in groups B and E. Because hydrogen peroxide concentrations in culture medium during the actual use of these dishes are lower than 0.5 µg/mL, representing the threshold for human mesenchymal stem cell viability [22], the µDrop GPS dishes in groups A, C, and D may exhibit strong or relatively high toxicity considering the residual amounts.

As shown in Fig 1, the mouse blastocyst rate in the control group was 85.7±6.7%, but the rate of the test group B was significantly decreased to 70.8±3.4% (p < 0.05). The rates of test group E recovered to 90.5±6.7%, reaching the same level as the control group, indicating that residual hydrogen peroxide was sufficiently removed to a level that had no effect on the blastocyst.

**Fig 1.**
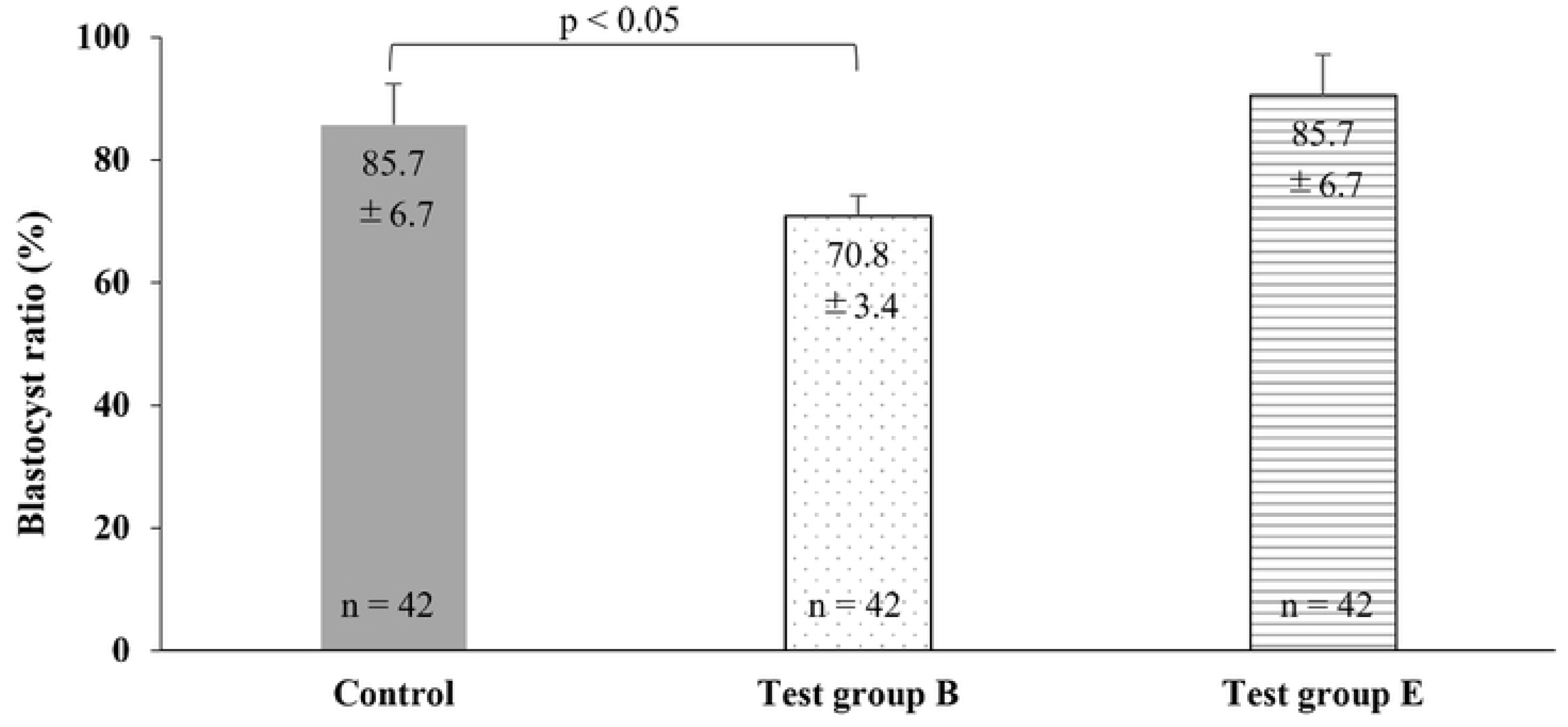
Effect of residual hydrogen peroxide on mouse embryo development. The control group, which used the commercial product without modification, was compared with the test groups B and E, as presented in Table 3. Values are the mean ± standard deviation.

Similar results were observed in HEA. As shown in Fig 2A, no significant difference was observed in the total development, including early blastocyst, between the control group and test group E. However, both full and good blastocyst ratios on day 5 in test group B were significantly reduced to 50.0±8.2 and 26.7±20.5% (p < 0.05), respectively, compared with those of the control group (Fig 2B), indicating that the sensitivity of human embryos to hydrogen peroxide is higher than that of mouse embryos.

**Fig 2.**
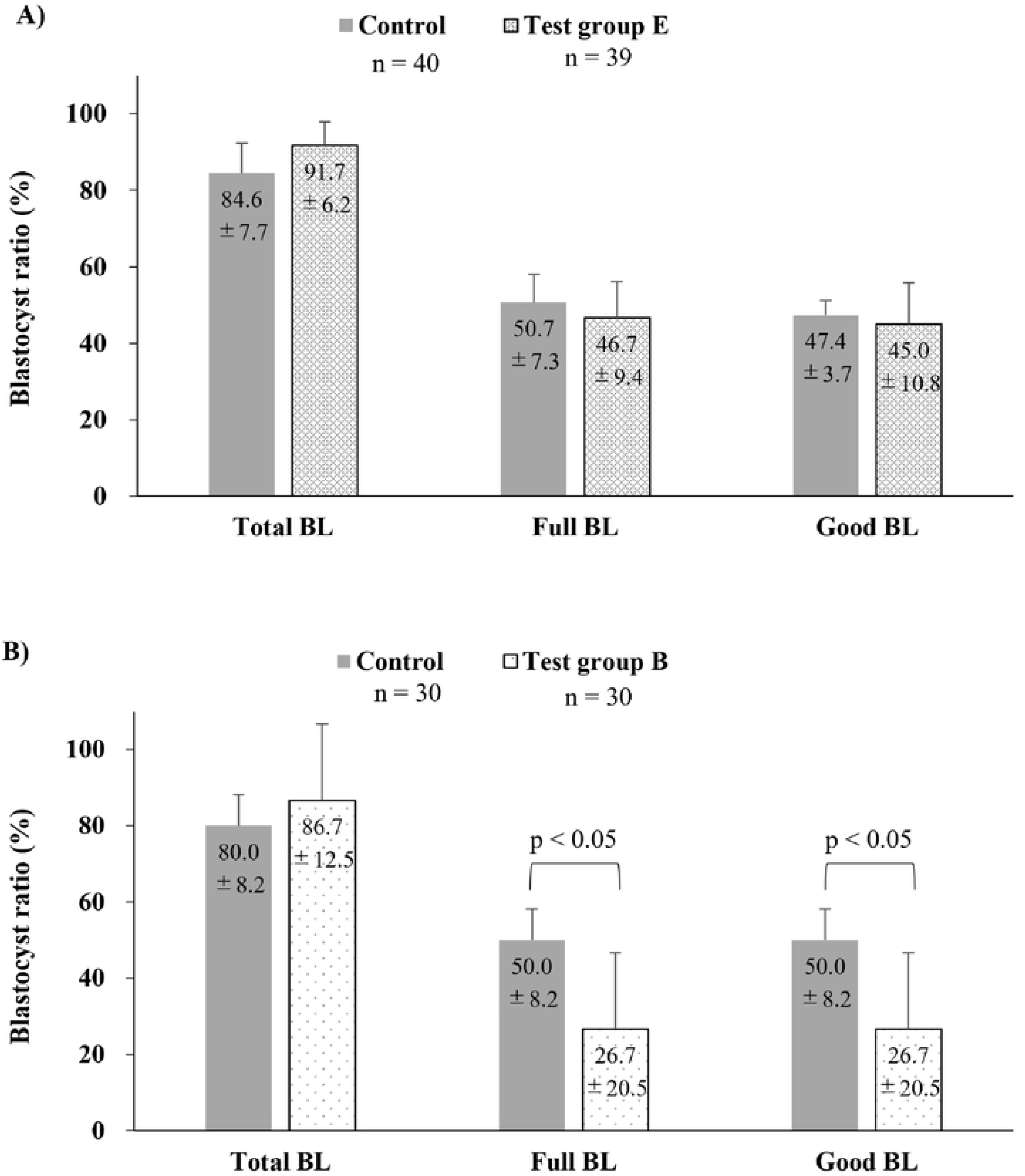
Effect of residual hydrogen peroxide on human embryo development. A) Blastocyst ratio at day 5 in control and test group E. B) Blastocyst ratio at day 5 in control and test group B. The control group, which used the commercial product without modification, was compared with the test groups B and E, as presented in Table 3. The blastocysts (BL) that had obtained a score of 3BB or more by using Gardner’s score [36] were defined as good BL. Values are the mean ± standard deviation.

### Effect on sperm survival test

The effect of residual hydrogen peroxide on the viability of human sperm was evaluated. As shown in Fig 3, the motility ratio of sperm in the control group cultured with a non-treated GPS dish was 93.4±1.4% initially, but it significantly decreased to 66.6±12.0 and 33.0±17.2% after 24 and 48 h of incubation, respectively. The GPS dish subjected to ozone/hydrogen peroxide gas treatment, followed by 455 aeration cycles in the sterilizer (test group B), showed cytotoxicity, and the motility ratio was greatly decreased to 16.2±15.3 and 4.1±4.4% after 24 and 48 h of culture, respectively (p < 0.01). On the other hand, the GPS dish treated with ozone/hydrogen peroxide gas sterilization, followed by drying treatment at 80℃ for 24 h (test group E), exhibited no toxicity, and the sperm survival curve was almost equal to that of the control group (motility ratio: 70.6±10.2 and 33.9±18.1% after 24 and 48 h culture).

**Fig 3.**
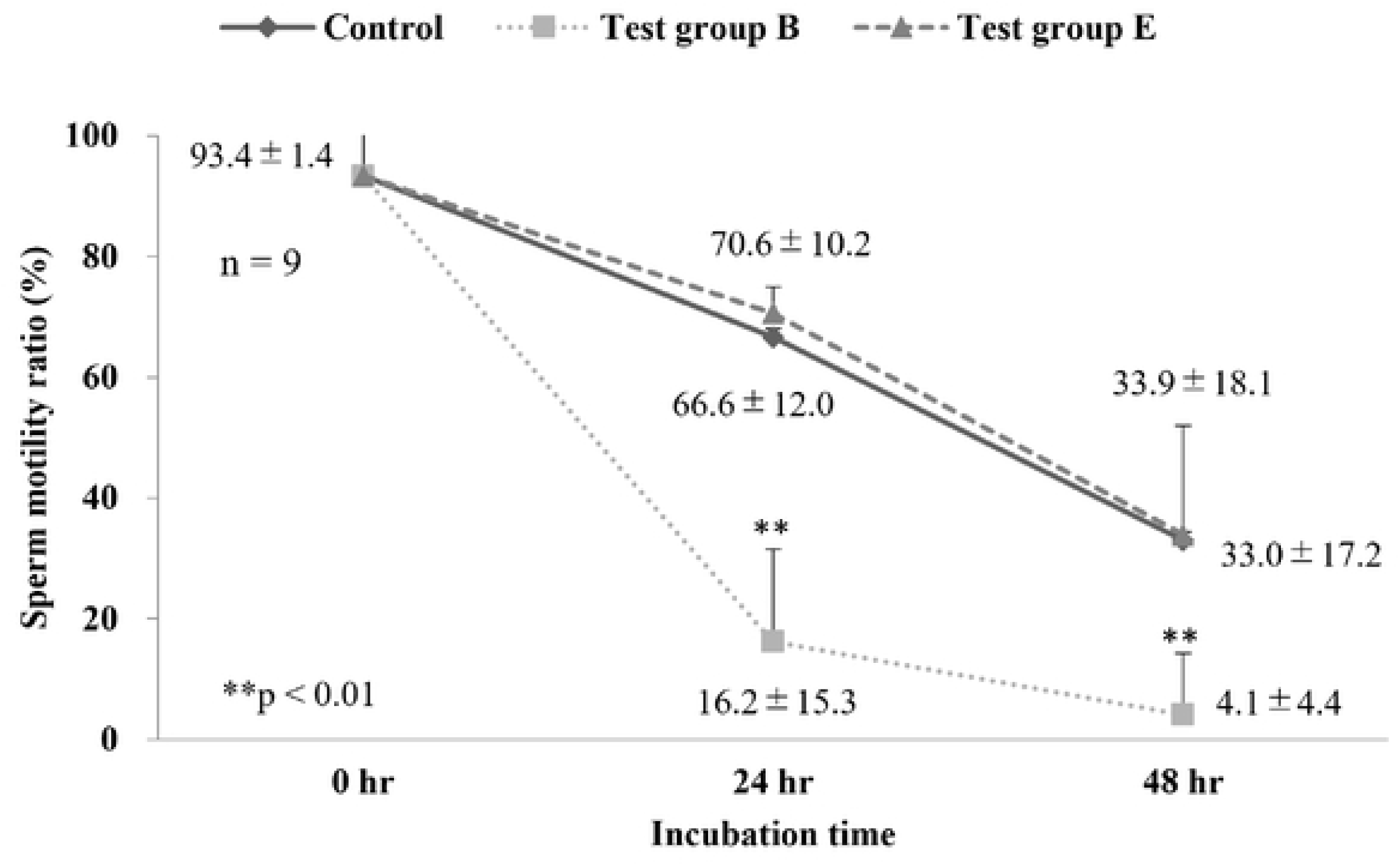
Effect of residual hydrogen peroxide on the viability of human sperm. Sperm motility ratio after incubation for 0 h, 24 h, and 48 h at 37°C under 5% CO_2_ and 5% O_2_. The control group, which used the commercial product without modification, was compared with the test groups B and E, as presented in Table 3. Values are the mean ± standard deviation.

## Discussion

Given that heat treatment at 250℃ for more than 30 min is required for complete inactivation of endotoxin [23], heat loads in a range of 150 to 180°C during the pelletizing and molding steps in the manufacturing process of plastic products are not sufficient for depyrogenation. Existing sterilization methods are not effective for reliable depyrogenation [24], and hence, commercially available plastic cell culture utensils are manufactured in an automated clean space to minimize pyrogen contamination and shipped after radiation sterilization [25]. In addition, since endotoxin is chemically a lipopolysaccharide with phosphate groups [4,5] and adsorbs to various materials via chelate, hydrophobic, and ionic bonding, the endotoxin test for materials, including plastic products, using water extracts carries a risk of false-negative results due to low recovery [16,17,19,20,26]. For these reasons, manufacturers of the utensils may overlook pyrogen contamination of their products, although the quality of the utensils used in experiments and product development should be high to obtain reproducible data and manufacture products with optimal safety [27,28].

HCPT is an in vitro pyrogen assay that measures the amounts of IL-6 released from immune cells as a fever marker. It is officially listed in the European Pharmacopoeia as the monocyte activation test (MAT), which is an alternative method to the pyrogen test using rabbits [29.30]. The United States Pharmacopoeia also allows evaluation by MAT if the equivalence is verified [31]. Although MAT is scheduled to be included in the nineteenth edition of the Japanese Pharmacopoeia, which will be revised in 2026, direct HCPT can also be used for biological safety evaluation of medical devices in Japan as a new method [17–20,26]. Unlike the endotoxin test, HCPT, including MAT, has the advantage of being unaffected by the recovery ratio, because relatively small-sized samples can be directly co-cultured with cells by immersion in the medium without preparing water extracts [17,18].

As shown in Table 1, samples nos. 5, 9, and 13 were labeled “Non-pyrogenic,” and nos. 1, 2, 4, 6 to 8, and 11 were set to the endotoxin levels according to the manufacturer’s own criteria. Sample nos. 15 to 20, as medical devices approved in Japan, may have endotoxin limits set by the manufacturer as one of the biological safety indicators. Although other samples were only labeled as “Sterile” or “DNA & Nuclease free,” these utensils used for germ cell collection and culture should basically be pyrogen-free. However, HCPT showed that seven samples were pyrogen-positive; particularly, pyrogen levels of nos. 5, 12, and 16 to 18 may exceed the endotoxin limit (2 pg/mL equal to 0.01-0.02 EU/mL) for human IVF [2,3] during actual use of the products. The use of these utensils, highly contaminated with pyrogen, may reduce ART success rates. In ART, even subtle environmental stressors can affect fertilization rates, blastocyst development, implantation success, and ultimately pregnancy and live birth outcomes [32]. Thus, pyrogen contamination represents not only a theoretical concern but also a clinically significant determinant of ART outcomes. The present study demonstrated that ozone/hydrogen peroxide sterilization in ET mode could decrease the pyrogen level of these utensils to below LOD. Most samples were able to inactivate pyrogens with a single ET mode treatment, but sample no. 17, with a relatively long silicone shaft, required three treatments to remove pyrogens. This phenomenon is thought to be caused by poor gas permeability, hydrogen peroxide adsorption, or both. In addition, ozone/hydrogen peroxide gas sterilization does not affect material properties such as Fourier Transform Infrared Spectroscopy spectra, static contact angle, and tensile strength of not only polypropylene and polystyrene [16], which correspond to raw materials for cell culture utensils, but also soft polyvinyl chloride, silicone, nylon, and polysulfone (data not shown), which comprise intrauterine insemination and embryo transfer catheters.

Thus, the sterilization method using the advanced oxidation process of ozone [19] is very useful to improve the cleanliness of the products at low temperature (50℃) and to more safely culture gametes and embryos in ART, but hydrogen peroxide remaining on the surface of the sterilized utensils is highly toxic to cells as an oxidative stress agent [22,33,34]. Reduction of blastocyst rate and sperm motility rate when cultured on µDrop GPS dishes of test group B may result from the amount of residual hydrogen peroxide (0.37±0.03 µg/mL), suggesting human germ cells may be slightly more sensitive to hydrogen peroxide than human mesenchymal stem cells, of which the toxicity threshold for viability is 0.5 µg/mL [22]. However, drying treatment at 80°C for 24 h after the sterilization was effective in eliminating residual hydrogen peroxide toxicity to human sperm culture and embryogenesis. This is extremely important in ensuring the safety of culture devices used in ART. In clinical practice, where the number of transferrable embryos is often limited, minimizing risks from contaminated devices is essential for successful pregnancy outcomes. Compared with other low-temperature sterilization methods, such as ethylene oxide gas sterilization and gamma ray sterilization, the ozone/hydrogen peroxide sterilization used in this study has the advantage of leaving fewer residues and having a high capacity for endotoxin inactivation at low temperatures, and it is expected that this method will become the standard method in the future.

Because the technology developed in this study to improve the quality of utensils used in human IVF may be difficult to introduce to individual hospitals, it is ideal that manufacturers use this technology to deliver highly clean products to users. The establishment of highly clean culture devices is expected to lead to the optimization of the embryo culture environment in ART and to improved clinical outcomes.

## Conclusion

Commercially available culture utensils and catheters currently used for human IVF may carry a potential risk of reducing ART success rates. However, the present study suggests that pathogen-free manufacturing of these products with no adverse effects on germ cells is now possible using ozone/hydrogen peroxide gas sterilization and subsequent drying technologies, improving ART success rates. Our clinic has replaced all materials used for IVF with pyrogen-free products. ART success rates in our clinic will be monitored and statistically compared with historical results.

## Data Availability

All relevant data are within the manuscript and its Supporting Information files.

## Acknowledgements

We would like to thank Editage (www.editage.jp) for English language editing.

## References

1. Japan Association of Obstetricians and Gynecologists. Assisted reproductive technology (ART), Obstetrics and Gynecology Seminar. 2025 [cited 2025 December 18]. Available from: https://www.jaog.or.jp/lecture/11-%E7%94%9F%E6%AE%96%E8%A3%9C%E5%8A%A9%E5%8C%BB%E7%99%82%EF%BC%88art%EF%BC%89//1000 [In Japanese]

2. Nagata Y, Shirakawa K. Setting standards for the levels of endotoxin in the embryo culture media of human in vitro fertilization and embryo transfer. Fertil Steril. 1996;65: 614–619. doi: 10.1016/S0015-0282(16)58164-3.

3. Nagata Y, Koga Y, Kikutake Y. Culture medium and endotoxin. Journal of Mammalian Ova Research. 2003;20: 51–54 [In Japanese]. doi: 10.1274/jmor.20.51.

4. Rietschel ET, Wollenweber HW, Zähringer U, Lüderitz O, Westphal O, Brade H. Bacterial lipopolysaccharides and their lipid A component. In: Homma JY, Kanegasaki S, Lüderitz O, Shiba T, Westphal O, editors, Bacterial endotoxin: Chemical, biological and clinical aspects. Weinheim: Wiley-VCH Verlag GmbH; 1984, pp. 11-22.

5. Rietschel ET, Schade U, Seydel U, Zähringer U, Lindner B. Morgan AP, et al. Chemical structure and biological activity of lipopolysaccharides. In: Baumgartner JB, Calandra T, Carlet J, editors. Endotoxin from pathophysiology to therapeutic approaches. Paris: Flammarion Medicine-Sciences; 1990, pp. 5-18.

6. Snyman E, Van der Merwe JV. Endotoxin-polluted medium in a human in vitro fertilization program. Fertil Steril. 1986;46: 273–276. doi: 10.1016/s0015-0282(16)49525-7.

7. Han Q, Ji X, Chang L, Jin X, Wang C, Li J. The embryo toxicity research of manufacturing water of medical device of in vitro fertilization technology. Chinese Journal of Medical Instrumentation. 2020;44: 439–442. doi: 10.3969/j.issn.1671-7104.2020.05.014.

8. Dumoulin J, Menheere P, Evers J, Kleukers A, Pietres M, Bras M, et al. The effects of endotoxins on gametes and preimplantation embryos cultured in vitro. Hum Reprod. 1991;6: 730–734. doi: 10.1093/oxfordjournals.humrep.a137417.

9. Akira S, Uematsu S, Takeuchi O. Pathogen recognition and innate immunity. Cell. 2006;124: 783-801. doi: 10.1016/j.cell.2006.02.015.

10. Eley A, Hosseinzadeh S, Hakimi H, Geary I, Pacey AA. Apoptosis of ejaculated human sperm is induced by co-incubation with Chlamydia trachomatis lipopolysaccharide. Hum Reprod. 2005;20: 2601–2607. doi: 10.1093/humrep/dei082.

11. Fujita Y, Mihara T, Okazaki T, Shitanaka M, Kushino R, Ikeda C, et al. Toll-like receptors (TLR) 2 and 4 on human sperm recognize bacterial endotoxins and mediate apoptosis. Hum Reprod. 2011;26: 2799–2806. doi: 10.1093/humrep/der234.

12. Kitchens RL, Thompson PA. Modulatory effects of sCD14 and LBP on LPS-host cell interactions. J Endotoxin Res. 2005;11: 225–229. doi: 10.1179/096805105X46565.

13. Malm J, Nordahl EA, Bjartell A, Sørensen OE, Frohm B, Dentener MK, et al. Lipopolysaccharide-binding protein is produced in the epididymis and associated with spermatozoa and prostasomes. J Reprod Immunol. 2005;66: 33–43. doi: 10.1016/j.jri.2005.01.005.

14. Romero R, Roslansky P, Oyarzun E, Wan M, Emamian M, Novitsky TJ, et al. Labor and infection. II. Bacterial endotoxin in amniotic fluid and its relationship to the onset of preterm labor. Am J Obstet Gynecol. 1988;158: 1044–1049. doi: 10.1016/0002-9378(88)90216-5.

15. Viana GA, Cela V, Ruggiero M, Pluchino N, Genazzani AR, Tantini C. Endometritis in infertile couples: The role of hysteroscopy and bacterial endotoxin. JBRA Assist Reprod. 2015;19: 21–23. doi: 10.5935/1518-0557.20150006.

16. Ibrahim S, Nakamura Y, Sugizaki E, Koyama G, Chan Z, Tamura S, et al. Development of pyrogen-free utensils for cell culture by combining ozone/hydrogen peroxide mixed gas sterilization with steam-washing technology. Regen Ther. 2025;30: 157–163. doi: 10.1016/j.reth.2025.05.015.

17. International Organization for Standardization. Pyrogenicity – Principles and methods for pyrogen testing of medical devices. ISO/TR 21582. 2021 [cited 2025 December 18]. Available from: https://www.iso.org/standard/71150.html

18. Japanese Ministry of Health, Labor and Welfare. Yakuseikishin‒hatsu 0106 No. 1. Basic principles of biological safety evaluation are required for application for approval to market medical devices. Attachment: guidance on test methods for biological safety evaluation of medical devices. Part 7: Pyrogen Test. 2020 Jan 6 [cited 2025 December 18]. Available from: https://www.pmda.go.jp/files/000233545.pdf [In Japanese]

19. Nomura Y, Yamamura J, Fukui C, Fujimaki H, Sakamoto K, Matsuo K, et al. Performance evaluation of bactericidal effect and endotoxin inactivation by low-temperature ozone/hydrogen peroxide mixed gas exposure. J Biomed Mater Res B Appl Biomater. 2021;109: 1807–1816. doi: 10.1002/jbm.b.34840.

20. Nomura Y, Fukui C, Yamamura J, Kuromatsu H, Naito T, Takahashi Y, et al. Evaluation of pyrogens remaining on reusable medical devices after washing and sterilization. Japanese Journal of Medical Instrumentation. 2021;91: 323–331. doi: 10.4286/jjmi.91.323.

21. Gardner DK, Lane M, Stevens J, Schlenker T, Schoolcraft WB. Blastocyst score affects implantation and pregnancy outcome: towards a single blastocyst transfer. Fertil Steril. 2000;73: 1155–1158. doi: 10.1016/s0015-0282(00)00518-5.

22. Chihara R, Kitajima H, Ogawa Y, Nakamura H, Tsutsui S, Mizutani M, et al. Effects of residual H_2_O_2_ on the growth of MSCs after decontamination. Regen Ther. 2018;9: 111–115. doi: 10.1016/j.reth.2018.08.003.

23. 23. Japanese Pharmacopoeia 18th edition. Section 4.01: Bacterial endotoxins test. 2021 [cited 2025 December 18]. Available from: https://www.mhlw.go.jp/content/11120000/000945683.pdf

24. Hosobuchi K, Tanamoto K. Inactivation of dry endotoxin by several sterilization methods. Bulletin of Tokyo Metropolitan Industrial Technology Research Institute. 1999;2: 126–129. Available from: https://www.iri-tokyo.jp/uploaded/attachment/319.pdf [In Japanese with English title and abstract]

25. European Comission. EU Guidelines for good manufacturing practice for medicinal products for human and veterinary use: The rules governing medicinal products in the European Union, Volume 4, Annex 1 Manufacture of sterile medicinal products. 2022 [cited 2025 December 18]. Available from: https://health.ec.europa.eu/system/files/2022-08/20220825_gmp-an1_en_0.pdf

26. Haishima Y, Murai T, Nakagawa Y, Hirata M, Yagami T, Nakamura A. Chemical and biological evaluation of endotoxin contamination on natural rubber latex products. J Biomed Mater Res. 2001;55: 424–432. doi: 10.1002/1097-4636(20010605)55:3<424::aid-jbm1032>3.0.co;2-s.

27. Japanese Ministry of Economy, Trade and Industry. Japan Agency for Medical Research and Development. Guidelines on Requirements for Process Materials Specific to Cell Processing 2017. 2017 [cited 2025 December 18]. Available from: https://www.meti.go.jp/policy/mono_info_service/healthcare/iryou/downloadfiles/pdf/201703.32. pdf [In Japanese]

28. Forum for Innovative Regenerative Medicine Supporting Industries Committee. Plastic Products in Regenerative Medicine. Seikei-Kakou. 2019;31: 420–422. doi: 10.4325/seikeikakou.31.420 [In Japanese]

29. Hasiwa M, Kullmann K, von Aulock S, Klein CL, Hartung T. An in vitro pyrogen safety test for immune-stimulating components on surfaces. Biomaterials. 2007;28: 1367–1375. doi: 10.1016/j.biomaterials.2006.11.016.

30. European Directorate for the Quality of Medicines & HealthCare. Monocyte activation test: European pharmacopoeia, 11th ed. Strasbourg: Council of Europe; 2023.

31. United States Pharmacopoeial Convention. 151. Pyrogen Test: United States Pharmacopoeia, 9th ed. Rockville: United States Pharmacopoeial Convention; 2022.

32. Castillo CM, Harper J, Roberts SA, O’Neill HC, Johnstone ED, Brison DR. The impact of selected embryo culture conditions on ART treatment cycle outcomes: a UK national study. Hum Reprod Open. 2020;10: hoz031. doi: 10.1093/hropen/hoz031

33. Bekeschus S, Liebelt G, Menz J, Singer D, Wende K, Schmidt A. Cell cycle-related genes associate with sensitivity to hydrogen peroxide-induced toxicity. Redox Biol. 2022;50: 102234. doi: 10.1016/j.redox.2022.10223.

34. Singer D, Miebach L, Bekeschus S. Differential sensitivity of two leukemia cell lines towards two major gas plasma products, hydrogen peroxide and hypochlorous acid. Applied Sciences. 2022;12: 7429. doi: 10.3390/app12157429

